# Magnitude of antenatal depression among women attending antenatal care at public health centers in post-war Shire, Tigray region, Ethiopia: a facility based cross-sectional study

**DOI:** 10.64898/2026.08.24.26361194

**Authors:** Daniel Berhane Gebremikael, Teklehaimanot Gereziher Haile, Wagnew Tesfaye Gebresilase, Yohans Tadese, Taddis Brhane

**Affiliations:** Tigray Health Research Institute, Tigray, Ethiopia; Department of Nursing, College of Medicine and Health Sciences, Aksum University, Tigray, Ethiopia; Tigray Health Research Institute, Tigray, Ethiopia.; Department of Public Health, College of Medicine and Health Sciences, Adigrat University, Tigray, Ethiopia; Department of Reproductive Health, College of Medicine and Health Sciences, Aksum University, Tigray, Ethiopia

**Keywords:** Antenatal depression, prenatal mental health, conflict, IDPs, Ethiopia

## Abstract

**Introduction:** Antenatal depression is a major public health concern linked to adverse maternal and neonatal outcomes, including preterm birth, impaired fetal growth, low birth weight, infant malnutrition, and increased episodes of childhood illness. This study assessed the magnitude and factors associated with antenatal depression among pregnant women attending public health centers in Shire town, Tigray, Ethiopia.

**Methods:** This facility-based cross-sectional study allocated the sample proportionally across health centers based on November–December 2025 antenatal care caseloads. After selecting the first participant by lottery, every third eligible attendee was enrolled through systematic sampling. Variables with p≤0.25 in bivariable analysis were entered into a multivariable logistic regression model to identify factors associated with antenatal depression among pregnant women in post-war Shire Town, Tigray, Ethiopia.

**Results:** All 463 participants were included (response rate: 100%). The magnitude of antenatal depression was 34.3% (95% CI: 30–38.7%). Increased odds of depression were observed among age group 25-34 years (AOR = 3.43; 95% CI: 1.7–7.2), those with unplanned pregnancies (AOR=2.2; 95% CI: 1.25–3.87), exposure to conflict-related traumatic events (AOR=2.6; 95% CI: 1.41–4.81), Internally displaced people (AOR=2.02; 95% CI: 1.05–3.92), experience of intimate partner violence (AOR=2.22; 95% CI: 1.31–3.8), poor partner relationship (AOR=2.17; 95% CI: 1.2-3.96), and low perceived neighborhood safety (AOR=2.7; 95% CI: 1.5–5.08). Protective factors included middle income (AOR=0.54;(95% CI:0.31-0.93), higher income (AOR=0.15; 95% CI: 0.065–0.33), and very good pre-war economic status (AOR=0.4; 95% CI: 0.16–0.94).

**Conclusion:** Antenatal depression was common among pregnant women in this post-war population, with internally displaced women experiencing higher odds. Integrating mental health and psychosocial support into antenatal care, alongside interventions addressing conflict-related trauma, intimate partner violence, socioeconomic vulnerability, and community safety, is warranted.

## Introduction

Pregnancy is a period characterized by substantial physical, psychological, social, and economic changes. While pregnancy is often regarded as a time of joy and anticipation, these transitions can also be stressful and emotionally demanding, heightening women’s vulnerability to mental health problems, particularly depression[1]. Women’s experience of pregnancy and other factors shape their attitude towards it; for some, it is a time of new experience and joy; for others, it is a stressful time, and depending on these and other intricate factors, throughout the world, 10 % of pregnant women experience a mental health disorder [2,3], and maternal depression during pregnancy remains a major public health concern, indicating the importance of addressing maternal depression [4].

Antenatal depression refers to a spectrum of non-psychotic depressive symptoms experienced during pregnancy, ranging from mild to severe [5–7]. According to the fifth edition of the Diagnostic and Statistical Manual of Mental Disorders (DSM-5), antenatal depression is classified as a major depressive disorder with peripartum onset, which is characterized by the occurrence of at least one major depressive episode in the absence of manic or hypomanic episodes [8].

The consequences of antenatal depression extend beyond maternal psychological well-being, imposing substantial economic burdens on families and societies. In Pakistan, untreated perinatal depression and anxiety were estimated to cost over 16.5 billion USD in 2017 (approximately 2,680 USD per birth) [9]. Similarly, women with postpartum depression in South Africa were more likely to experience unemployment, lower household income, and socioeconomic disadvantage than those without depression [10].

Antenatal depression is associated with adverse maternal, fetal, and infant health outcomes. Infants born to mothers with antenatal depression have a 49% higher risk of low birth weight and a 40% higher risk of preterm birth [4,11]. Maternal depression has also been linked to increased odds of preterm birth, low birth weight, childhood malnutrition, and acute febrile illness, as well as stillbirth, perinatal complications, postpartum depression, and impaired fetal growth [4].

Given these significant maternal, fetal, infant, and socioeconomic consequences, early identification and management of antenatal depression have become important components of quality maternal healthcare. Routine screening for depression during ANC provides an opportunity to identify affected women and initiate appropriate interventions. The impact of antenatal depression may be particularly pronounced in conflict-affected settings where displacement, socioeconomic hardship, disruption of health services, and exposure to traumatic events increase vulnerability to mental health disorders.

Evidence suggests that antenatal depression is common globally, although prevalence estimates vary considerably across populations and measurement tools [4,12]. Despite its well-documented adverse maternal and infant health consequences, antenatal depression remains under-recognized and under-detected in many settings. Similarly, experiencing prenatal exposure to natural disasters aggravates the chance of having antenatal depression [13]. Untreated maternal depression can contribute to elevated risks of stillbirth, obstetric complications, operative delivery, and postpartum depression [4,11,14,15].

Studies conducted at low and middle-income countries estimate the prevalence of antenatal depression (AND) to be between 15.6 and 25.3 %[6,12]. Studies from LMICs identify several predictors of antenatal depression, including childhood sexual abuse, intimate partner violence, unintended pregnancy, low socioeconomic status, limited social support, a history of mental illness, low educational attainment, being unmarried, young maternal age, and exposure to disasters[6,12,16]. Conversely, protective factors include higher education, supportive intimate partnerships, and stable employment[16].

National studies on AND prevalence conducted using different validated tools indicate a wide variation, but an umbrella study indicated 24.6% AND prevalence[5], while a meta-analysis and systematic review also indicated closer values, i.e., 24.24% [1]. Ethiopian studies consistently identify socioeconomic disadvantage, poor social support, intimate partner violence, adverse reproductive experiences, and a history of mental illness as major determinants of antenatal depression[1,17–24].

Before the Tigray war, a facility-based study conducted among 196 pregnant women using the Beck Depression Inventory (BDI) reported an antenatal depression prevalence of 31.1%, with being unmarried and being a housewife identified as significant predictors [25]. This study was conducted before the war, and studies conducted after the war show a decline in maternal healthcare utilization and conditions compared to prewar[26]. Reduced ANC utilization may further limit opportunities for depression screening and management among pregnant women in post-conflict settings. The war in Tigray has had profound consequences on maternal mental health and healthcare access. Evidence from conflict-affected settings indicates that war-related financial instability, loss of social support, and heightened concerns about child well-being significantly exacerbate maternal psychological distress [27]. Studies among general internally displaced persons (IDPs) in Tigray found depression prevalence as high as 81.2%, demonstrating the extreme mental health burden in post-conflict populations[28]. However, no study has examined the burden of antenatal depression specifically among pregnant women attending ANC services in post-war Tigray, particularly in Shire town.

The findings of this study have important implications for achieving the United Nations Sustainable Development Goals (SDGs). The high prevalence of antenatal depression and its association with conflict-related experiences, internal displacement, low household income, inadequate social support, and low perceived neighborhood safety highlight the need to integrate maternal mental health screening and care into routine antenatal services, alongside psychosocial and social support interventions. These actions directly support SDG 3 (Good Health and Well-being), particularly Target 3.4 on promoting mental health and well-being, while indirectly contributing to improved maternal health under Target 3.1. The disproportionate burden among internally displaced women also underscores the need to reduce health inequalities (SDG 10) and promote gender-responsive maternal healthcare (SDG 5). Furthermore, the associations with conflict exposure, poverty, and neighborhood safety emphasize the importance of advancing SDG 16 (Peace, Justice and Strong Institutions), SDG 1 (No Poverty; Targets 1.4 and 1.5), and SDG 11 (Sustainable Cities and Communities) to address the broader social determinants of maternal mental health[29].

Maternal depression during pregnancy is a critical yet under-recognized public health issue with substantial implications for maternal well-being, fetal development, and neonatal outcomes. In post-war settings such as Shire town, psychological distress is likely to be exacerbated by displacement, loss of livelihoods, weakened social support, and disrupted health services. However, evidence on the magnitude of antenatal depression in post-war Tigray remains scarce, limiting efforts to design targeted screening, early detection, and intervention strategies within antenatal care (ANC).

This study provides timely, context-specific evidence on the prevalence of antenatal depression among pregnant women attending ANC in public health centers in Shire. The findings can inform post-war recovery efforts, strengthen maternal health services, and improve maternal and child health outcomes. Future longitudinal studies are needed to clarify causal pathways and examine the impact of antenatal depression on birth outcomes while accounting for broader social and institutional factors.

The absence of post-war, context-specific evidence on antenatal depression in Shire and the wider Tigray region remains a major barrier to designing effective maternal mental health interventions and integrating routine depression screening into ANC services. Local evidence is essential to support evidence-based public health planning, guide equitable resource allocation, and strengthen advocacy for integrating mental health care within maternal health programs. Therefore, this study was conducted to determine the magnitude of antenatal depression and identify associated factors among pregnant women attending ANC services in public health facilities in Shire town, Tigray, Ethiopia.

## Methods and materials

### Study design and Setting

A facility-based cross-sectional study was conducted among pregnant women attending antenatal care services at Shire town, in the Northwest zone of Tigray, Ethiopia, from March 4-30/2026.

#### Source population

All pregnant women who attend antenatal care (ANC) services at governmental public health centers in Shire city.

#### Study sample

A subset of the study population consisting of pregnant women attending ANC services at governmental public health centers in Shire city during the study period who meet the eligibility criteria and are selected for inclusion in the study.

#### Study participants

Pregnant women from the study sample who provided informed consent and completed the study.

### Eligibility criteria

#### Inclusion criteria

All pregnant women aged 18 and above who visited the antenatal care (ANC) clinics at Alganesh Health Center, Umer Health Center, and Five Angeles Health Center have been included.

#### Exclusion criteria

Women who were in labour, severely ill pregnant women, and people who had trouble communicating throughout the data collection time were all excluded from this study.

### Sample size determination

A single population proportion formula was used to determine the sample size. Sample size calculation was performed using OpenEpi software. A 95% confidence interval (CI) and a margin of error (d) of 4% were used. This level of precision is consistent with standard practice for facility-based cross-sectional studies estimating prevalence and has been widely applied in previous Ethiopian studies assessing the magnitude of antenatal depression and its associated factors [17,18,20,21].

Sample size was also calculated for the main determinant variables expected to have a strong association with the outcome using -OpenEpi Software. The variable that yielded the largest sample size was considered as the final sample size for the study.

**Table 1:** Magnitude of antenatal depression among women attending antenatal care at public health centers in post-war Shire, Tigray region, Ethiopia: a facility based cross-sectional study; Sample size determination from primary and secondary objectives.

| Parameter selected | Reference citation | P1 and q1 values | Sample size |
| --- | --- | --- | --- |
| Magnitude of AND In Ethiopia-proportion of AND | [5] | P1: 24.6%, Q1:75.4 | 446 |
| Proportion of Women with exposure to Intimate Partner Violence-IPV | [21] | P1:20.9 q1: 79.1 | 397 |
| Proportion of women with has got a social support | [20] | P1:29.9% q1:70.1% | 503 |
| Proportion of women who said ‘their partner has done things to scare or intimidate them (IPV) | [20] | P1:16.9% q1: 83.1% | 338 |
| Proportion of women who are single and with AND | [20] | P1:79%, q1: 21% | 399 |

The prevalence of social support from friends reported in the study “Antenatal Depression and Associated Factors among Pregnant Women Attending Antenatal Care Service in Kochi Health Center, Jimma Town, Ethiopia, 2020” was used to estimate the initial sample size using OpenEpi, yielding 503 participants.

Because the source population was finite and known, and the initial sample size represented a substantial proportion (20.9%) of the expected ANC population of 2,400, the finite population correction (FPC) was applied to avoid overestimation and ensure efficient sampling. The corrected sample size was 416.

The total source population was based on the two-month ANC load (November–December 2025) across the three public health centers in Shire:

- Alganesh Health Center (Na) = 1,400 (58.3%)
- Umer Health Center (Nu) = 600 (25.0%)
- Five Angels Health Center (Nf) = 400 (16.7%)
- Total population (N) = 2,400

To account for a 10% non-response rate, the final sample size was calculated as:

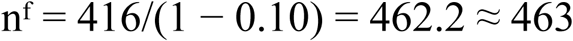

Thus, the final sample size was 463 participants.

Proportional allocation of the sample was performed according to the ANC load of each health centre:

- Alganesh Health Center: (1,400/2,400) × 463 = 270
- Umer Health Center: (600/2,400) × 463 = 116
- Five Angels Health Center: (400/2,400) × 463 = 77

Therefore, 270, 116, and 77 participants were allocated to Alganesh, Umer, and Five Angels Health Centers, respectively.

### Sampling technique and procedure

All public health centers in Shire town were included in the study, namely Alganesh Health Center, Umer Health Center, and Five Angles Health Center. The total sample size was determined based on the proportion of pregnant women with good social support reported in a previous study conducted at Kochi Health Center, Jimma, using ANC attendance from the preceding two months as the reference period. The final sample size was proportionally allocated to each health center according to their ANC client volume recorded during November and December 2025, with a total of 2,400 ANC visits across the three facilities. Within each health center, study participants were selected using a systematic random sampling technique.

The sampling interval (k) for each facility was calculated by dividing the expected number of ANC clients during the one-month data collection period by the sample size allocated to that facility. ANC client flow estimates were taken based on November and December service statistics. All interviews were conducted at ANC exit points.

The first participant in each facility was selected randomly using the lottery method from the expected eligible ANC clients attending within the first two days of data collection. Subsequently, every kth eligible pregnant woman who consents to participate was interviewed until the allocated sample size for each facility is attained.

Facility-specific kth interval calculation:

`o` Alganesh Health Center: 700/270 = 3
`o` Umer Health Center: 300/116 = 3
`o` Five Angles Health Center: 200/77 = 3

The expected ANC caseload over 44 working days (November–December), for Alganesh HC was 1,400, corresponding to approximately 700 clients per month. A total of 184 participants will be sampled from Alganesh Health Center over a month. (Fig 1)

**Fig 2:**
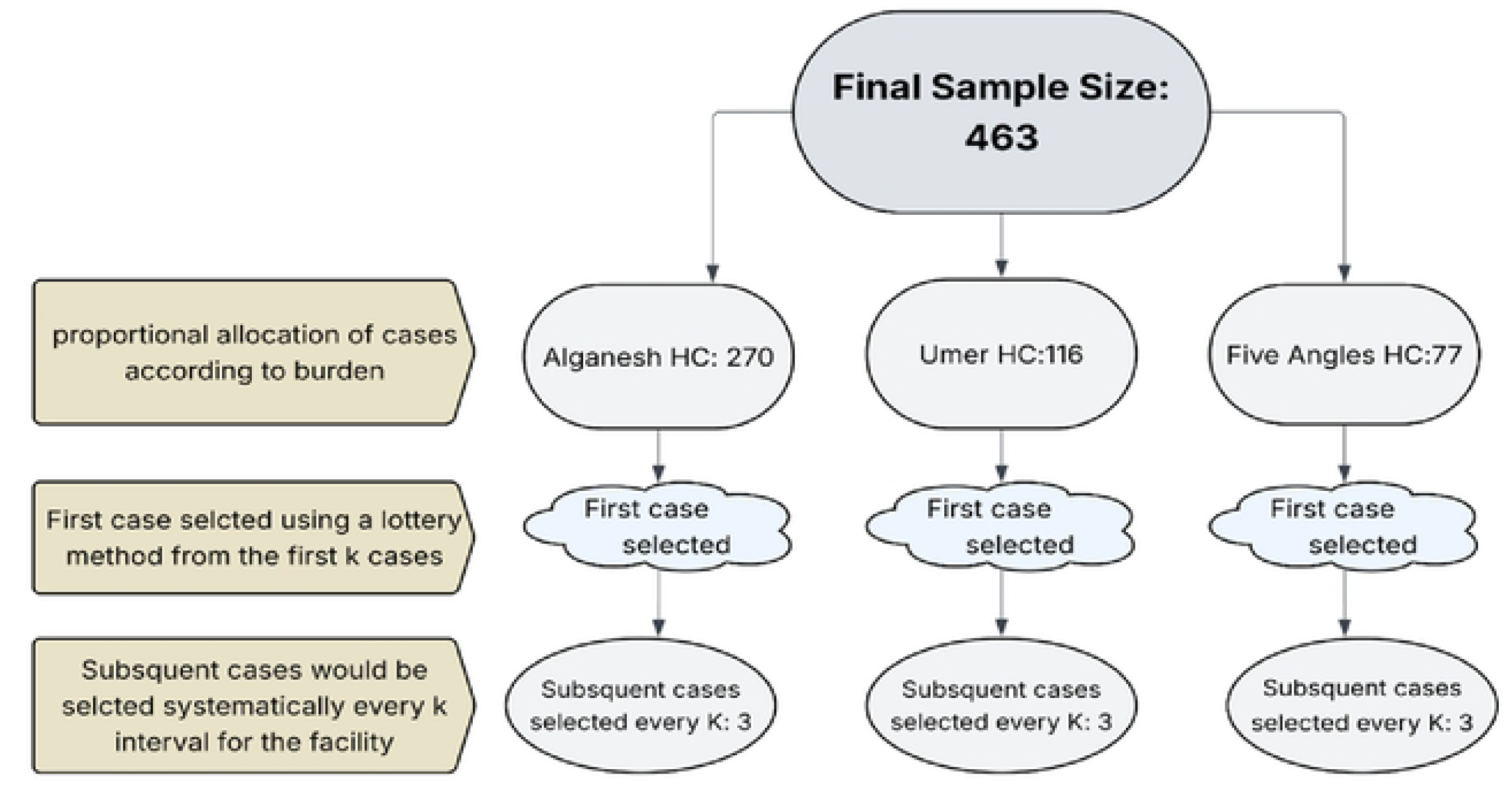
Sampling procedure.

**Fig 2:**
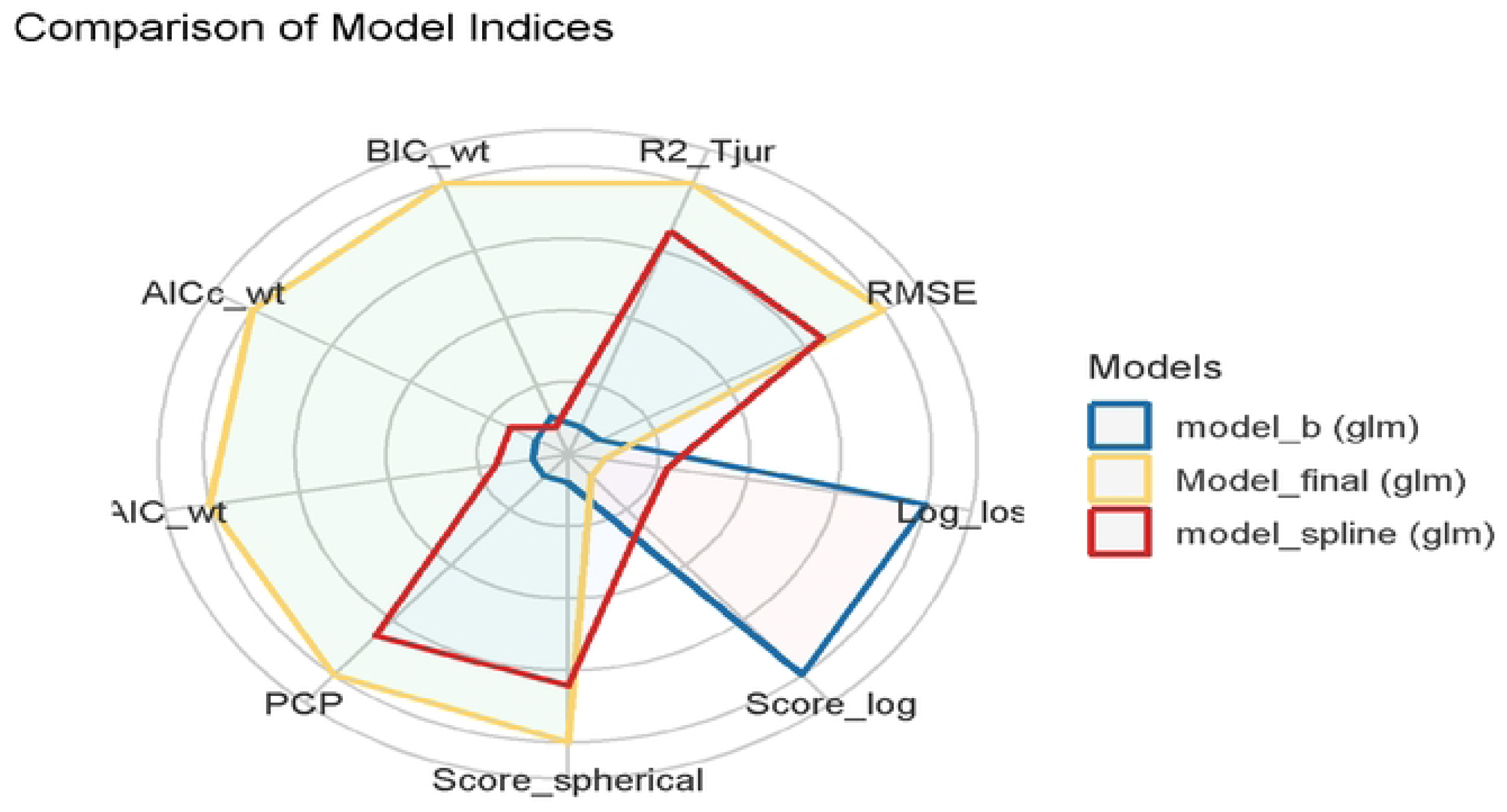
Model performance comparison.

### Variables

#### Dependent Variable

Antenatal depression was assessed using the nine-item Patient Health Questionnaire (PHQ-9) among women selected according to the predefined eligibility criteria and facility-specific Kth interval through systematic random sampling. Women who scored greater than or equal to 10 (≥10) were classified as having depression, whereas those who scored less than 10 (<10) were classified as not having depression. Each of the nine PHQ-9 items is scored on a scale ranging from 0 to 3, resulting in a total possible score ranging from 0 to 27.

#### Independent Variable

`o` Individual: age, educational status, parity, occupation, income, gestational age in trimesters, substance use, pregnancy planned, experienced complications during pregnancy, experienced complication during current pregnancy, known chronic medical illness, has diagnosed mental health problem, family history of mental health problem, displacement status, lost family member, experienced conflict related event, prewar economic status, post war economic status.
`o` Interpersonal: partner relationship quality, partner emotional support, Intimate partner violence (IPV).
`o` Community: perceived neighborhood safety score -PNSS.
`o` Institutional: ANC service quality, ANC counselling service quality

### Operational Definitions

**Age:** Age in years was categorized into three categories. Age less than 24 years, 25- 34 years, and ≥ 35 years. **Antenatal Depression:** Antenatal depression was measured using the Patient Health Questionnaire-9 (PHQ-9); a total score of ≥10 was classified as antenatal depression (“Yes”), while a score <10 was classified as “No.”

**ANC service quality:** Assessed by asking mothers to rate the quality of ANC services received during their current visit using a 5-point Likert scale: very poor = 1, poor = 2, fair = 3, good = 4, excellent = 5. Responses were categorized as **poor quality** (very poor, poor, fair) and **good quality** (good and excellent).

**ANC counselling service quality:** Assessed by asking mothers to rate the quality of counselling services received during their antenatal care visit using a 5-point Likert scale: very poor = 1, poor = 2, fair = 3, good = 4, excellent = 5. Responses were categorized as **poor quality** (very poor, poor, fair) and **good quality** (good and excellent).

**Current pregnancy complication:** Participants selected one or more complications from a list (severe nausea/vomiting, vaginal bleeding, high blood pressure or preeclampsia, gestational diabetes, pregnancy loss or preterm labour, ectopic pregnancy, others) or selected “no complication.” Any reported complication was classified as “Positive complication,” otherwise “No complication.”

**Chronic medical illness:** Presence of chronic medical illness (e.g., DM, hypertension, HIV/AIDS). Classified as “Yes” if present and “No” if absent.

**Intimate partner violence (IPV):** The Abuse Assessment Screen (AAS) was used to assess IPV. A “Yes” response to any of the questions 1–5 of the five-item AAS indicated IPV (“Yes”).

**Educational status:** Participants are asked about their highest attained educational status, and their responses are labelled into no formal education and primary education (grades 1–8), which in turn were collapsed into “primary or no education”, and Secondary education (grades 9–12) and college diploma or above were retained as separate categories.

**Experienced conflict-related event:** Exposure to traumatic events such as violence, loss of property, or displacement was classified as “Yes,” otherwise “No.”

**Experienced complication during previous pregnancy:** Participants selected from a list of complications (severe nausea/vomiting, vaginal bleeding, high blood pressure or preeclampsia, gestational diabetes, pregnancy loss or preterm labour, ectopic pregnancy, others) or “no complication” and “Not applicable” was also used for nulliparous women. Any reported complication was classified as “Positive complication,” otherwise “No complication,” or “Not applicable” was selected as per the participant’s response.

**Gestational age (trimester):** First trimester (≤13 weeks), second trimester (14–27 weeks), and third trimester (≥28 weeks).

**Income (Ethiopian birr):** Income was categorized using the 25th percentile (4,000 birr) and 75th percentile (10,000 birr) into three categories: <4,000 = low income, 4,000–10,000 = middle income, and >10,000 = high income.

**Intimate Partner Violence:** The Abuse Assessment Screen (AAS) was used to assess and measure IPV. A “Yes” response to any of the questions 1–5 of the AAS was classified as IPV (“Yes”).

**Known chronic medical illness (DM, hypertension, HIV/AIDS, etc.):** Classified as “Yes” if present and “No” if absent.

**Marital status:** Respondents reporting “married” were classified as “married,” while divorced, widowed, or cohabiting were grouped as “non-married.”

**Occupation:** Government employees, farmers, and other categories were collapsed into “other,” while housewives and merchants were retained as separate categories.

**Parity:** Nulliparous = no previous birth, primiparous = one previous birth, multiparous = more than one previous birth.

**Partner relationship quality:** Participants are asked to grade their relationship quality with their partner in a 5-point Likert scale. Rated as excellent-5, good-4, fair-3, poor-2, or very poor-1. Responses were categorized as **good** (excellent and good) and **poor** (fair, poor, very poor).

**Partner emotional support:** Participants are asked if they feel emotionally supported by their partner during this pregnancy, and grade it in a Likert scale. Response options included always-5, often-4, sometimes-3, rarely-2, and never-1. **Adequate support** included always and often, while **inadequate support** included sometimes, rarely, and never.

**Pregnancy intention/planned/:** Classified as “planned” if the pregnancy was intended, otherwise “unplanned.”

**Perceived neighbourhood safety scale (MESA study):** Assessed using a three-item scale measuring safety while walking, absence of violence, and perceived safety from crime. Items were scored from 1 (strongly agree-1, agree-2, neutral-3, disagree-4, strongly disagree-5), with total scores ranging from 3 to 15. A score ≥9 indicated low perceived neighbourhood safety, while a score <9 indicated high perceived neighbourhood safety.

**Prewar economic status:** Assessed by the question: “How would you describe your household socioeconomic status before the war?” Response options included very good-4, good enough-3, not very bad-2, very bad-1, and not applicable-0. Very good was categorized as “**Very Good**,” while good enough, not very bad, and very bad were categorized as “**Not bad**,” and **not applicable** was retained.

**Postwar economic status:** Assessed by the question: “How has your household socioeconomic status changed after the war?” Response options included Much better-5, improved-4, unchanged-3, somewhat worsened-2, much worsened-1, and not applicable-0. Much better, improved, and unchanged were categorized as “**Not deteriorated**,” while somewhat worsened and much worsened were categorized as “**Deteriorated**,” and **not applicable** was retained.

**Substance use/Alcohol use/:** Classified as “Yes” if the participant used one or more of alcohol, khat, or cigarettes, and “No” if none were used. In this study, ‘Substance use’ can also be used interchangeably as ‘alcohol use,’ as all women with a “Yes” response used alcohol only, except one case who concomitantly used ‘Khat’.

### Data collection tool and procedure

Structured questionnaires that had been validated and pretested were used to collect data from the study participants through face-to-face interviews. Following a 2- day training led by the primary investigator, the data were collected by two BSc community health professionals, and a follow-up training was given after 2 weeks of data collection as well to ensure mutual learning and address issues. On the second day of training, the data collectors were trained on the Kobo interface familiarization, and they practiced with each other as a role-play drama on the actual content of the questionnaire.

### Data quality control

Pretest was done at Endabaguna primary hospital, from February 10-13/2026. Submitted questionnaires were checked for consistency and completeness daily. High priority was given to the design of data gathering instruments to ensure data quality, and ensuring Kobo’s inbuilt error checking mechanisms, before submission of data was put as a priority during data collection. The questionnaire was prepared in English, translated into Tigrinya (the local language) by fluent speakers of the language, and back-translated to English by another person to maintain consistency. Information was gathered via questionnaires that were translated into Tigrinya and encoded into Kobo XLSForms.

Feedback on the daily submitted data was shared daily, and daily monitoring of the data collection process was conducted by the principal investigator.

### Performance of Tools Used

Description and measurements of construct measurement tools used in this research:

#### Reliability test

The MESA Neighborhood Safety scale demonstrated moderate internal consistency (Cronbach’s α = 0.57; McDonald’s ω = 0.61). Although these values fall below the conventional 0.70 threshold, this level of reliability is not uncommon for brief three-item scales. The PHQ-9 showed acceptable internal consistency (Cronbach’s α = 0.70; McDonald’s ω = 0.80).

### Data processing and analysis

Data were coded and analyzed using R software version 2026.04.0+526. The following R packages were used for data management, analysis, and model diagnostics: *pacman*, *tidyverse*, *readr*, *psych*, *e1071*, *nortest*, *car*, *splines*, *detectseparation*, *broom*, *ResourceSelection*, *pscl*, *pROC*, *performance*, *gtsummary*, *janitor*, *brglm2*, and *rms.* Descriptive statistics, including means, medians, frequencies, percentages, and standard deviations, were computed to summarize the data.

#### Checking assumptions for Binary logistic regression

Assumptions for binary logistic regression were assessed before model fitting. Multicollinearity was evaluated using variance inflation factors (VIF), with all values below 3, indicating no evidence of problematic multicollinearity. The linearity of the logit for continuous variables was assessed using restricted spline functions, which showed no meaningful deviation from linearity. Comparison of the spline model (AIC = 488.34; BIC = 608.33) with the final model (AIC = 484.01; BIC = 599.86) favored the more parsimonious final model. Independence of observations was ensured by including each participant only once.

#### Model Fitness Assessment

Model fit was assessed using the likelihood ratio test, which showed that the final model fit the data significantly better than the intercept-only model (χ² = 167.66, p < 0.001). The Hosmer–Lemeshow goodness-of-fit test indicated adequate calibration (χ² = 13.39, df = 8, p = 0.099), while the area under the receiver operating characteristic curve (AUC = 0.84) demonstrated good discriminative ability. The model showed moderate explanatory power (McFadden R² = 0.281; Nagelkerke R² = 0.420). Overall, the model demonstrated good fit, acceptable calibration, moderate explanatory power, and good discrimination. Adjusted odds ratios (AORs) with 95% confidence intervals and corresponding p-values are presented. (Fig 1).

Model comparison plot (Final model vs Spline). model_final(model for the current research), Model_spline (similar with model_final but age is: ns(age,df=3)), and model_b (which is similar with model_final except it categorizes age as a continuous variable).

## Results

A total of 463 participants were included in the study, and the non-response rate was zero.

### A: Individual Characteristics

The mean age was 27.0 years (SD = 5.44; SE = 0.25; 95% CI: 26.5–27.5), and the median age was 26 years (IQR = 8 years). The majority of participants were married (87.2%), and among women who screened positive for depression, 81.1% were married. Regarding educational status, 44.0% had attained secondary education, 37.1% had no formal or primary education, and 14.7% had college-level education or higher. Most participants were housewives (69.8%), followed by merchants (11.0%) and others (5.8%).

In terms of income, 48.4% of participants were in the interquartile range (4,000–10,000 birr), 28.7% were in the lowest quartile (<4,000 birr), and 22.9% were in the highest quartile (>10,000 birr). Among women with depression (n = 159), 40.9% were in the lowest income, 50.3% were in the middle income, and 8.8% were in the highest income.

More than half of the participants were multiparous (54.4%), while 37.4% were nulliparous. Regarding gestational age, 47.1% were in the third trimester, 43.4% in the second trimester, and 9.5% in the first trimester. The majority (74.5%) reported planned pregnancies, while 25.5% reported unplanned pregnancies. Regarding obstetric history, 50.8% reported no previous pregnancy complications, 18.1% reported at least one complication, and 31.1% were not applicable. For the current pregnancy, 83.8% reported no complications, while 16.2% had experienced at least one complication. Only 2.1% (n = 10) of participants reported a known chronic medical illness, while 97.9% reported none. A family history of mental illness was reported by 19.0%, and 2.8% had a personal history of diagnosed mental illness.

Regarding substance use during the current pregnancy, 64.1% reported use of at least one substance (alcohol, khat, or cigarettes), while 35.9% reported no use. Among women with depression, 74.2% reported substance use compared to 25.8% who did not. (Table 3)

**Table 2:**
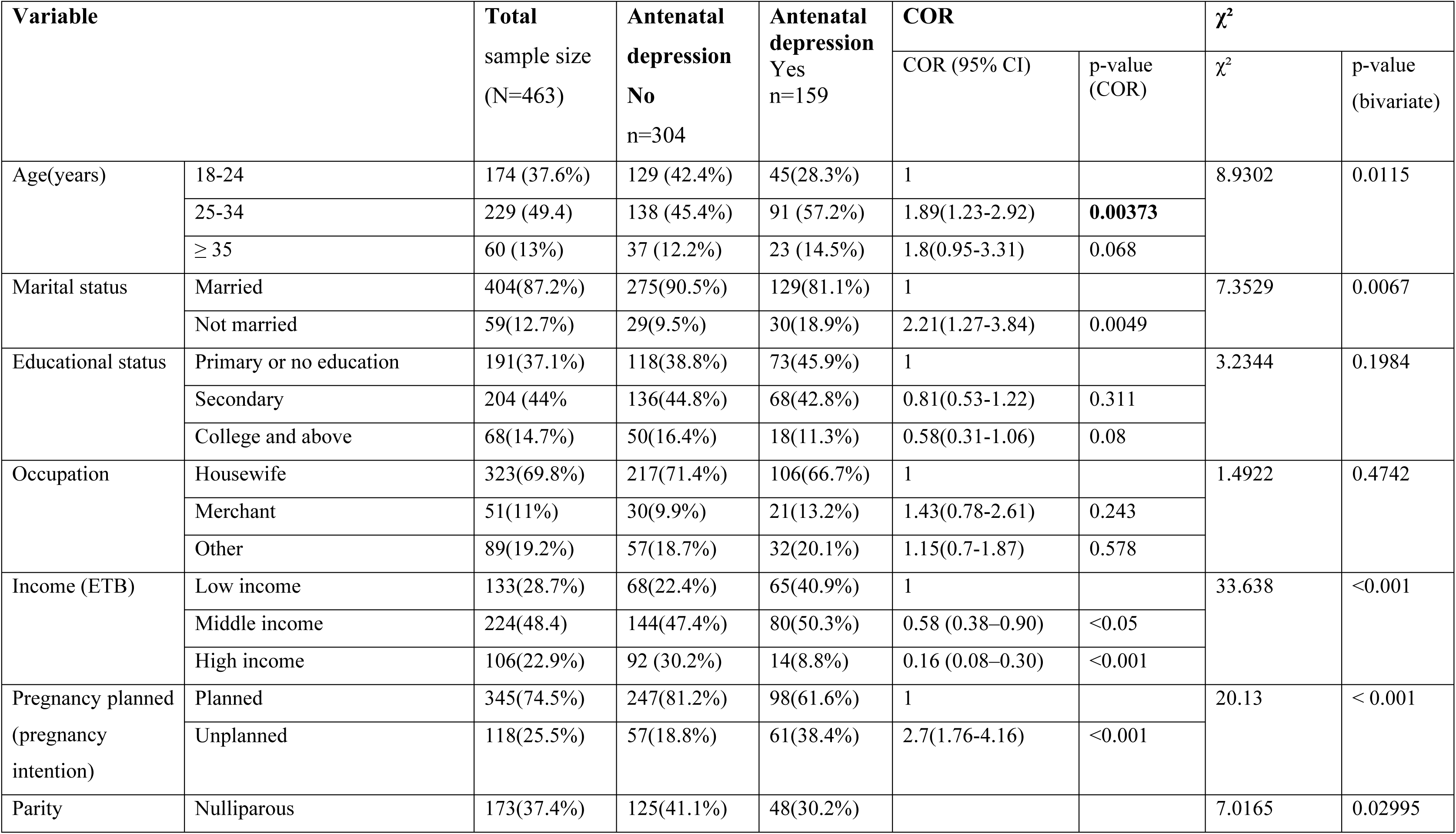
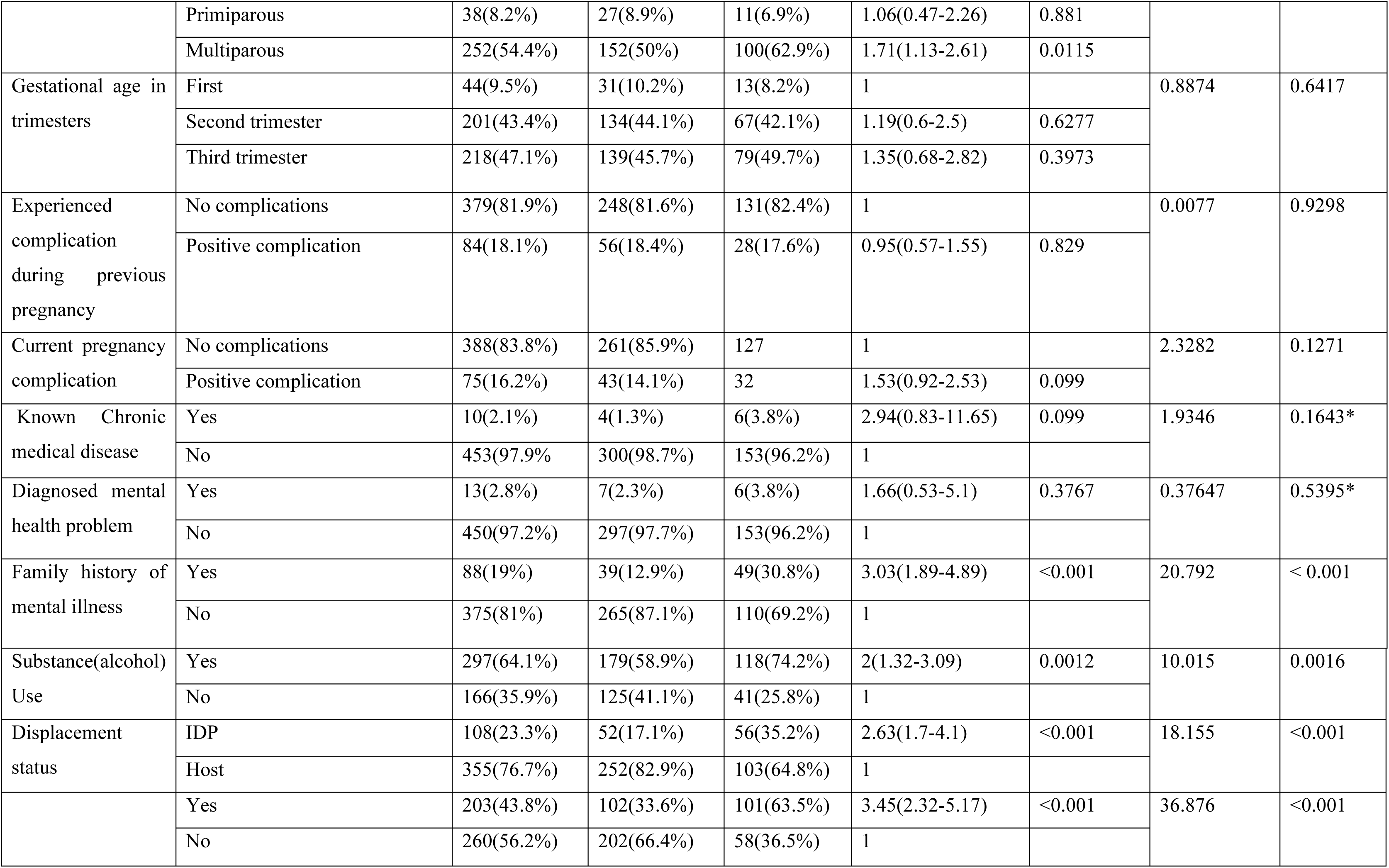
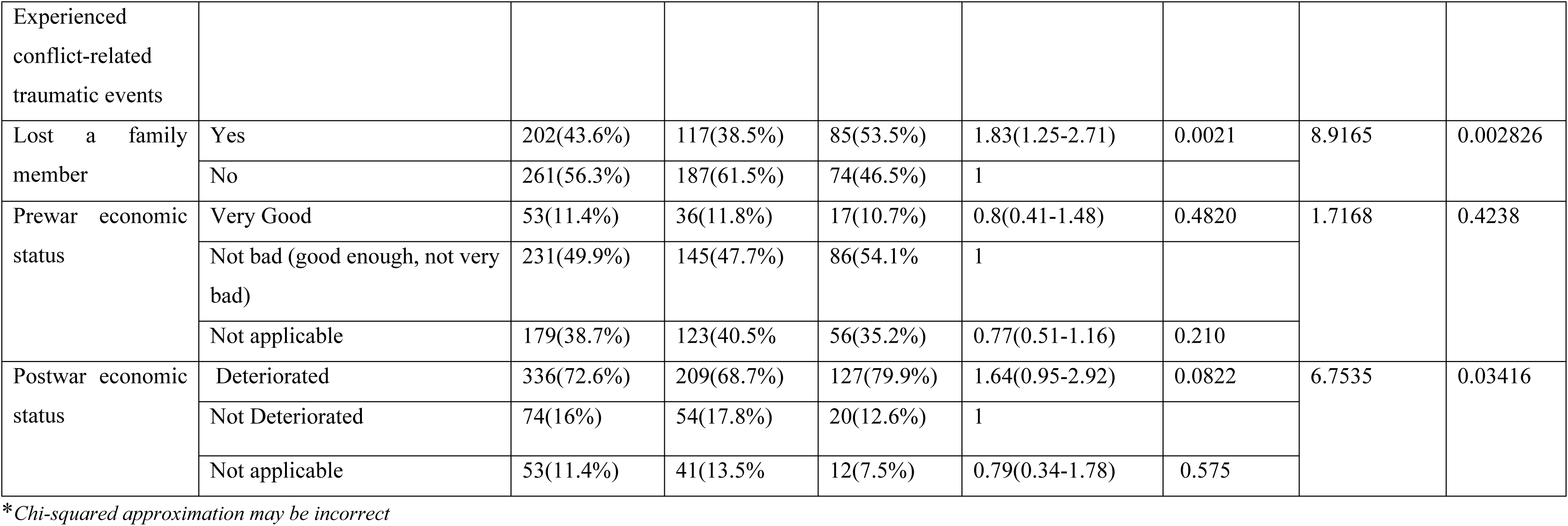
Magnitude of antenatal depression among women attending antenatal care at public health centers in post-war Shire, Tigray region, Ethiopia: a facility based cross-sectional study; Individual factors.

**Table 3:**
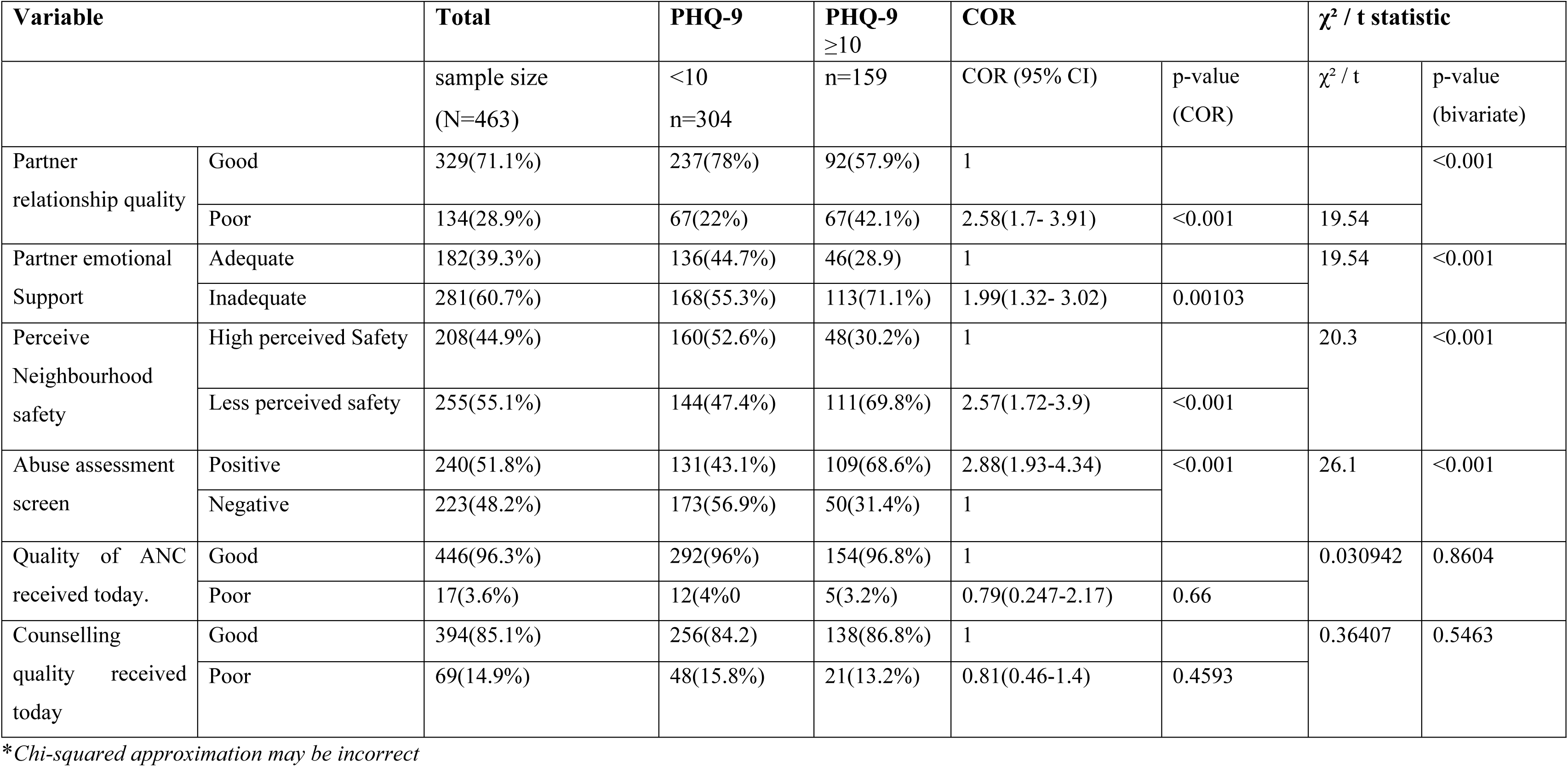
Magnitude of antenatal depression among women attending antenatal care at public health centers in post-war Shire, Tigray region, Ethiopia: a facility based cross-sectional study; Interpersonal, community, and institutional factors.

| | | sample size<br>(N=463) | <10<br>n=304 | n=159 | COR (95% CI) | p-value<br>(COR) | $\chi^2$ / t | p-value<br>(bivariate) |
| --- | --- | --- | --- | --- | --- | --- | --- | --- |
| Partner<br>relationship quality | Good | 329(71.1%) | 237(78%) | 92(57.9%) | 1 |  |  | <0.001 |
|  | Poor | 134(28.9%) | 67(22%) | 67(42.1%) | 2.58(1.7- 3.91) | <0.001 | 19.54 |  |
| Partner emotional<br>Support | Adequate | 182(39.3%) | 136(44.7%) | 46(28.9) | 1 |  | 19.54 | <0.001 |
|  | Inadequate | 281(60.7%) | 168(55.3%) | 113(71.1%) | 1.99(1.32- 3.02) | 0.00103 |  |  |
| Perceive<br>Neighbourhood<br>safety | High perceived Safety | 208(44.9%) | 160(52.6%) | 48(30.2%) | 1 |  | 20.3 | <0.001 |
|  | Less perceived safety | 255(55.1%) | 144(47.4%) | 111(69.8%) | 2.57(1.72-3.9) | <0.001 |  |  |
| Abuse assessment<br>screen | Positive | 240(51.8%) | 131(43.1%) | 109(68.6%) | 2.88(1.93-4.34) | <0.001 | 26.1 | <0.001 |
|  | Negative | 223(48.2%) | 173(56.9%) | 50(31.4%) | 1 |  |  |  |
| Quality of ANC<br>received today. | Good | 446(96.3%) | 292(96%) | 154(96.8%) | 1 |  | 0.030942 | 0.8604 |
|  | Poor | 17(3.6%) | 12(4%0 | 5(3.2%) | 0.79(0.247-2.17) | 0.66 |  |  |
| Counselling<br>quality received<br>today | Good | 394(85.1%) | 256(84.2) | 138(86.8%) | 1 |  | 0.36407 | 0.5463 |
|  | Poor | 69(14.9%) | 48(15.8%) | 21(13.2%) | 0.81(0.46-1.4) | 0.4593 |  |  |

\*Chi-squared approximation may be incorrect

The majority of participants (76.7%) were from host communities, while 23.3% were internally displaced persons (IDPs). Nearly half (43.8%) reported exposure to conflict-related events, and 43.6% reported loss of at least one family member.

Regarding economic conditions, 11.4% reported very good pre-war economic status, 49.9% reported “not bad” conditions, and the remainder were not applicable. Post-war, 72.6% reported no deterioration in economic status, while 16.0% reported deterioration.

### B: Interpersonal Factors

Most participants (71.1%) reported a good partner relationship, while 28.9% reported poor relationship quality. However, only 39.3% reported adequate emotional support from their partner during pregnancy, whereas 60.7% reported inadequate support. A positive screening for intimate partner violence was reported by 51.8% of participants, while 48.2% screened negative.

### C: Community Factors

Perceived neighborhood safety was assessed using a three-item scale adapted from the MESA Neighbourhood Safety Scale. The scale showed suboptimal but acceptable internal consistency (Cronbach’s α = 0.57; McDonald’s ω = 0.61) for a three-item scale. Overall, 44.9% of participants reported high perceived safety, while 55.1% reported low perceived safety.

### D. Institutional Factors

The majority of participants (96.3%) rated the quality of ANC services received during the interview visit as good, while 3.6% rated it as poor. Similarly, 85.1% rated the quality of counselling services as good, compared to 14.9% who rated it as poor.

### E. Magnitude of antenatal depression

The magnitude of antenatal depression among women attending antenatal care was 34.3%. (Table 3)

## BIVARIATE ANALYSIS

### Chi-square Test

A bivariate analysis (look at Table 3 and Table 4), using the chi-square test, was conducted to examine the association between categorical independent variables and antenatal depression.

**Table 4:** Magnitude of antenatal depression among women attending antenatal care at public health centers in post-war Shire, Tigray region, Ethiopia: a facility based cross-sectional study; Multivariable regression.

| Variable |  | Total sample size (N=463) | PHQ-9 <10 n=304 | PHQ-9 ≥10 n=159 | AOR |  |
| --- | --- | --- | --- | --- | --- | --- |
|  |  |  |  |  | AOR (95% CI) | p-value |
| Age(years) | 18-24 | 174 (37.6%) | 129 (42.4%) | 45(28.3%) |  |  |
|  | 25-34 | 229 (49.4) | 138 (45.4%) | 91 (57.2%) | 3.43(1.7-7.2) | **0.000776** |
|  | ≥ 35 | 60 (13%) | 37 (12.2%) | 23 (14.5%) | 2.13(0.8-5.71) | 0.131 |
| Marital status | Married | 404(87.2%) | 275(90.5%) | 129(81.1%) | 1 |  |
|  | Not married | 59(12.7%) | 29(9.5%) | 30(18.9%) | 0.7(0.31-1.6) | 0.404 |
| Educational status | Primary/no education | 191(37.1%) | 118(38.8%) | 73(45.9%) | 1 |  |
|  | Secondary | 204 (44%) | 136(44.8%) | 68(42.8%) | 1.02(0.6-1.76) | 0.944 |
|  | College or above | 68(14.7%) | 50(16.4%) | 18(11.3%) | 0.51(0.21-1.16) | 0.112 |
| Occupation | Housewife | 323(69.8%) | 217(71.4%) | 106(66.7%) | 1 |  |
|  | Merchant | 51(11%) | 30(9.9%) | 21(13.2%) | 1.51(0.68-3.32) | 0.307 |
|  | Other | 89(5.8%) | 57(18.7%) | 32(20.1%) | 1.5(0.75-2.86) | 0.267 |
| Income (ETB) | Low income | 133(28.7%) | 68(22.4%) | 65(40.9%) | 1 |  |
|  | Middle income | 224(48.4) | 144(47.4%) | 80(50.3%) | 0.54(0.31-0.93) | **0.0278** |
|  | High income | 106(22.9%) | 92 (30.2%) | 14(8.8%) | 0.15(0.065-0.33) | **0.000007** |
| Pregnancy planned (pregnancy intention) | Planned | 345(74.5%) | 247(81.2%) | 98(61.6%) | 1 |  |
|  | Unplanned | 118(25.5%) | 57(18.8%) | 61(38.4%) | 2.2(1.25-3.87) | **0.0062** |
| Parity | Nulliparous | 173(37.4%) | 125(41.1%) | 48(30.2%) | 1 |  |
|  | Primiparous | 38(8.2%) | 27(8.9%) | 11(6.9%) | 1.82(0.59-5.44) | 0.291 |
|  | Multiparous | 252(54.4%) | 152(50%) | 100(62.9%) | 1.83(0.85-4.01) | 0.127 |
| Prewar economic status | Very good | 53(11.4%) | 36(11.8%) | 17(10.7%) | 0.4(0.16-0.94) | **0.0387** |
|  | Not bad | 231(49.9%) | 145(47.7%) | 86(54.1%) | 1 |  |
|  | Not applicable | 179(38.7%) | 123(40.5%) | 56(35.2%) | 3.1(1.4-7.1) | **0.00644** |
| Current pregnancy complication | No complications | 388(83.8%) | 261(85.9%) | 127 | 1 |  |
|  | Positive complication | 75(16.2%) | 43(14.1%) | 32 | 1.57(0.81-3.03) | 0.1809 |
| Known Chronic medical disease | Yes | 10(2.1%) | 4(1.3%) | 6(3.8%) | 4.6(0.82-28.1) | 0.087 |
|  | No | 453(97.9%) | 300(98.7%) | 153(96.2%) | 1 |  |
| Family history of mental illness | Yes | 88(19%) | 39(12.9%) | 49(30.8%) | 1.8(0.98-3.2) | 0.056 |
|  | No | 375(81%) | 265(87.1%) | 110(69.2%) | 1 |  |
| Substance Use | Yes | 297(64.1%) | 179(58.9%) | 118(74.2%) | 1.52(0.81-2.89) | 0.194 |
|  | No | 166(35.9%) | 125(41.1%) | 41(25.8%) | 1 |  |
| Displacement status | IDP | 108(23.3%) | 52(17.1%) | 56(35.2%) | 2.02(1.05-3.92) | **0.037** |
|  | Host | 355(76.7%) | 252(82.9%) | 103(64.8%) | 1 |  |
| Experienced conflict-related traumatic events | Yes | 203(43.8%) | 102(33.6%) | 101(63.5%) | 2.6(1.41-4.81) | **0.00233** |
|  | No | 260(56.2%) | 202(66.4%) | 58(36.5%) | 1 |  |
| Lost a family member | Yes | 202(43.6%) | 117(38.5%) | 85(53.5%) | 1.5(0.91-2.48) | 0.114 |
|  | No | 261(56.3%) | 187(61.5%) | 74(46.5%) | 1 |  |
| Postwar economic status | Not Deteriorated | 336(72.6%) | 209(68.7%) | 127(79.9%) | 1 |  |
|  | Deteriorated | 74(16%) | 54(17.8%) | 20(12.6%) | 0.68(0.33-1.39) | 0.279 |
|  | Not applicable | 53(11.4%) | 41(13.5%) | 12(7.5%) | 1.3(0.42-3.93) | 0.644 |
| Partner relationship quality | Good | 329(71.1%) | 237(78%) | 92(57.9%) | 1 |  |
|  | Poor | 134(28.9%) | 67(22%) | 67(42.1%) | 2.17(1.2-3.96) | **0.0108** |
| Partner emotional support | Adequate | 182(39.3%) | 136(44.7%) | 46(28.9) | 1 |  |
|  | Inadequate | 281(60.7%) | 168(55.3%) | 113(71.1%) | 0.94(0.54-1.67) | 0.8433 |
| Perceive neighbourhood safety | High perceived Safety | 208(44.9%) | 160(52.6%) | 48(30.2%) | 1 |  |
|  | Less perceived safety | 255(55.1%) | 144(47.4%) | 111(69.8%) | 2.7(1.5-5.08) | **0.00133** |
| Abuse assessment screen | Positive | 240(51.8%) | 131(43.1%) | 109(68.6%) | 2.22(1.31-3.8) | **0.00313** |
|  | Negative | 223(48.2%) | 173(56.9%) | 50(31.4%) | 1 |  |

Age, marital status, monthly income, pregnancy planning status, parity, family history of mental illness, substance use, displacement status, exposure to conflict, loss of a family member, partner relationship quality, partner emotional support, perceived neighborhood safety, intimate partner violence (as measured by the Abuse Assessment Screen), and post-war economic status were significantly associated with antenatal depression (p < 0.05).

In contrast, no statistically significant associations were observed for educational status, occupation, gestational age (trimester), previous pregnancy complications, current pregnancy complications, known chronic medical illness, diagnosed mental health conditions, pre-war economic status, quality of ANC services, or counselling quality.

### Simple logistic regression

Bivariate logistic regression analysis was performed to assess the crude association between independent variables and antenatal depression.

Strong statistically significant associations (p < 0.001) were observed for several factors. Women with high monthly income (>10,000 ETB) had significantly lower odds of depression (COR = 0.16; 95% CI: 0.08–0.30). In contrast, higher odds of depression were observed among women with unplanned pregnancies (COR = 2.70; 95% CI: 1.76–4.16), a family history of mental illness (COR = 3.03; 95% CI: 1.89–4.89), displacement (COR = 2.63; 95% CI: 1.70–4.10), exposure to conflict (COR = 3.45; 95% CI: 2.32–5.17), poor partner relationship quality (COR = 2.58; 95% CI: 1.70–3.91), low perceived neighborhood safety (COR = 2.57; 95% CI: 1.72– 3.90), and intimate partner violence (COR = 2.88; 95% CI: 1.93–4.34).

Statistically significant associations at the 5% level were also observed for age category 25-34 years (COR=1.89, CI: 1.23-2.92), not being married (COR = 2.20; 95% CI: 1.27–3.84), middle income (COR = 0.58; 95% CI: 0.38–0.90), multiparity (COR = 1.71; 95% CI: 1.13–2.61), substance use (COR = 2.00; 95% CI: 1.32–3.09), loss of a family member (COR = 1.83; 95% CI: 1.25–2.71), and inadequate partner emotional support (COR = 1.99; 95% CI: 1.32–3.02).

No statistically significant associations (p > 0.05) were observed between antenatal depression and educational status, occupation, gestational age, previous pregnancy complications, current pregnancy complications, chronic medical illness, diagnosed mental health conditions, pre-war economic status, post-war economic status, ANC service quality, or counselling quality.

Variables with p ≤ 0.25 in the bivariate analysis were included in the multivariable logistic regression model.

### Multivariate Analysis

A multivariable binary logistic regression analysis was conducted to identify factors independently associated with antenatal depression after adjusting for potential confounders. (Table 5)

At the individual level, income showed a strong inverse association with depression. Compared to women in the low-income group, those with middle income had significantly lower odds of depression (AOR = 0.54; 95% CI: 0.31–0.93; p = 0.028), and women with high income had substantially reduced odds (AOR = 0.15; 95% CI: 0.065–0.33; p < 0.001).

Age was also associated with depression. Women aged 25–34 years had significantly higher odds compared to those aged <25 years (AOR = 3.43; 95% CI: 1.7–7.2; p < 0.001), while those aged ≥35 years showed increased but non-significant odds (AOR = 2.13; 95% CI: 0.8–5.71; p = 0.131).

Pregnancy intention remained an important factor, with women reporting unplanned pregnancies having higher odds of depression compared to those with planned pregnancies (AOR = 2.2; 95% CI: 1.25–3.87; p = 0.006).

Pre-war economic status showed differential associations. Women reporting a “very good” pre-war economic status had significantly lower odds of depression compared to those reporting “not bad” status (AOR = 0.40; 95% CI: 0.16–0.94; p = 0.039), while those in the “not applicable” category had significantly higher odds (AOR = 3.1; 95% CI: 1.4–7.1; p = 0.006).

Conflict-related factors remained strong predictors. Women who experienced conflict-related traumatic events had significantly higher odds of depression (AOR = 2.6; 95% CI: 1.41–4.81; p = 0.002), and internally displaced women had increased odds compared to host community participants (AOR = 2.02; 95% CI: 1.05–3.92; p = 0.037).

At the interpersonal level, exposure to intimate partner violence, as measured by the Abuse Assessment Screen, was significantly associated with higher odds of depression (AOR = 2.22; 95% CI: 1.31–3.8; p = 0.003). Poor partner relationship quality was also independently associated with increased odds of depression (AOR = 2.17; 95% CI: 1.2–3.96; p = 0.011).

At the community level, low perceived neighborhood safety was significantly associated with higher odds of depression (AOR = 2.7; 95% CI: 1.5–5.08; p = 0.001).

Several variables that were significant in the bivariate analysis, including marital status, occupation, substance use, partner emotional support, educational status, current pregnancy complications, chronic medical illness, family history of mental illness, parity, loss of a family member, and post-war economic status were not significantly associated with depression, were not significantly associated with depression after adjustment. Look at Table 5.

Institutional-level variables were not retained in the final multivariable model.

## Discussion

The present study aimed to assess the magnitude of antenatal depression and examine the effect of individual, interpersonal, community, and institutional-level factors among pregnant women attending antenatal care clinics in three public health centers in Shire town. The magnitude of antenatal depression among women attending antenatal care was 34.3%. This estimate is higher than the pooled AND prevalence reported in Ethiopia (24.6%) from a systematic review and meta-analysis [28], indicating greater burden in the post-war setting. Among internally displaced women in this study, the proportion of women with antenatal depression were 51.8%. This is lower than the prevalence of moderate to severe depression reported among community-based internally displaced persons during the Tigray war period (60%) [28]. However, this comparison should be interpreted with caution, as the previous study was conducted among a general displaced population during active conflict and not among pregnant women attending health facilities.

To our knowledge, this is among the first studies conducted in Shire city of Tigray region, following the recent Tigray war, after the cessation of hostilities agreement between the Federal Democratic Republic of Ethiopia (FDRE) and Tigray People’s Liberation Front (TPLF) [30], providing context-specific evidence from a post-war and displacement-affected population on antenatal depression.

In the multivariable analysis, after adjusting for potential confounders, several factors remained significantly associated with antenatal depression. These included lower income level, pregnancy intention (pregnancy plan), parity, exposure to conflict-related events, internal displacement status, pre-war economic status, intimate partner violence, poor partner relationship quality, and low perceived neighborhood safety.

The present study found an AND magnitude of 34.3%; this estimate exceeds pooled national and LMIC estimates (∼24–25%)[1,5,12]. This higher prevalence may reflect contextual differences, including socioeconomic instability, potential exposure to conflict-related stressors, sociocultural differences, and variations in study design and measurement tools, such as the use of tools, e.g., Patient Health Questionnaire (PHQ-9), Edinburgh Postnatal Depression Scale (EPDS), Beck Depression Inventory (BDI), or others. However, the observed prevalence remains within the global range of antenatal depression (15–65%) reported by an umbrella review[4], suggesting consistency with the wide variability documented across diverse settings.

In this study, women aged 25–34 years had significantly higher odds of antenatal depression compared with those aged 18–24 years (AOR = 3.43). A study conducted at Mubende Regional Referral Hospital also reported a significant association between maternal age and depression, particularly among women aged <20 years and ≥35 years.[31]. Although direct comparison is difficult because the age categories differed across the two studies, both findings suggest that maternal age may influence the likelihood of depression during pregnancy. In contrast, studies conducted in other parts of Ethiopia, like the Southern Ethiopia Study, the Gida Ayana District Western Ethiopia Study, and a study from comprehensive specialized hospitals in Northwest Ethiopia, reported no significant association between maternal age and antenatal depression[17–19]. One possible explanation for the observed higher odds of antenatal depression among women aged 25–34 years in the present study is the interplay between maternal age, parity, and caregiving responsibilities. Women aged 18–24 years are more likely to be primiparous or have fewer young children, resulting in a comparatively lower caregiving burden. In contrast, women aged 25–34 years often have larger families and multiple young children who still require substantial care and supervision but are generally not yet old enough to contribute meaningfully to household tasks or sibling care. Consequently, women in this age group may experience greater physical, emotional, and economic demands during pregnancy, particularly in post-war settings where family support systems may have been disrupted. By comparison, women at older maternal ages may have children who are sufficiently mature to assist with domestic work and caregiving responsibilities, potentially reducing maternal workload. Evidence from Ethiopia suggests that children’s contributions to household labour and caregiving increase with age, particularly among girls, although this potential mechanism was not directly assessed in the present study and should therefore be interpreted with caution[32,33].

Higher income (≥75th percentile, ≥10,000 birr) was significantly protective: women in this group had 84% lower odds of antenatal depression compared with those in the lowest-income category. Similarly, a facility-based study from Babile district of Eastern Ethiopia found lower income to have an increased odds of depression (AOR: 3.85) compared to higher income (>2000 birr)[34]. The protective effect of higher income may operate through several pathways, including increased income being associated with improved access to food security, better access to health care, reduced uncertainty, and greater coping resources. In contrast, a facility-based cross-sectional study from southern Ethiopia[18] and another from Uganda[31], both using the PHQ-9, reported no significant association between income and depression. Although conducted in a general population, a study from Northeast Ethiopia during the early phase of the Tigray War found no significant association between monthly income and depression, in contrast to our findings[35]. These discrepancies may partly reflect the unique socioeconomic conditions in post-war Tigray, where the slow recovery following the Cessation of Hostilities Agreement, persistent displacement, disruption of livelihoods, and reduced household purchasing power may have amplified the influence of household income on maternal mental health. In such settings, financial resources may be particularly important because they enable households to secure essential needs, including adequate nutrition, transportation to health services, and stable living conditions despite ongoing economic instability.

Unplanned current pregnancy was associated with nearly twofold higher odds of antenatal depression in this study (AOR = 1.96) compared with planned pregnancy, a finding that aligns with much of the existing evidence from Ethiopia. For instance, a facility-based study conducted in West Ethiopia (Gidey Ayana public health facilities)[17] reported a similar association (AOR = 2.07), and a systematic review and meta-analysis synthesizing 30 studies found that unplanned pregnancy was associated with substantially increased odds of depression (AOR = 3.04) relative to planned pregnancy[1]. Similarly, the previously cited Ugandan study also found a significant association with increased odds of depression among women with unplanned pregnancy [31]. The observed association is biologically and psychosocially plausible, as an unplanned pregnancy may increase psychological distress through reduced emotional preparedness for pregnancy, concerns about financial and caregiving responsibilities, and uncertainty regarding future family support.

However, some studies have reported contrasting findings. The previously cited study from southern Ethiopia did not observe a statistically significant association between unplanned pregnancy and depression[18]. Similarly, a study conducted in specialized hospitals in Northwest Ethiopia, which examined a related construct (unwanted pregnancy), found no significant relationship with depression[19]. These inconsistencies may be attributable to differences in how pregnancy intention is defined and measured, as well as variations in cultural perceptions of unintended pregnancy, partner and family support, and the socioeconomic and security conditions of the study populations. In the present post-war setting, these contextual factors may be particularly important, as war-related displacement, family separation, economic hardship, and disruption of social support networks could intensify the psychological burden associated with an unplanned pregnancy.

Having a “very good” prewar economic status was associated with 61% lower odds of depression compared with those reporting a “not bad” prewar status. This suggests that favorable pre-war economic conditions may confer resilience by supporting baseline food security, asset ownership, and coping capacity during and after periods of instability. Consistent with this interpretation, a study conducted seven years after conflict in Northern Uganda found that food insecurity was significantly associated with increased odds of major depressive disorder, and that female sex was also an independent risk factor[36]. However, prewar economic status in this study was assessed retrospectively and may be subject to recall and misclassification bias. In addition, individuals in post-war settings may reappraise prior living conditions more favourably relative to current hardship, a cognitive process often described as cognitive reappraisal, which could influence how past economic status is reported[37].

Being an internally displaced person (IDP) was associated with significantly higher odds of antenatal depression (AOR=2.07) compared with being from host community. This finding suggests that displacement-related vulnerabilities substantially increase psychological distress during pregnancy. Although conducted among IDPs living with community members, a study conducted in the Tigray Region indicates that IDPs, particularly those who have lost fixed assets, experienced crop looting and faced severe livelihood disruption, have a higher likelihood of depression than those who do not experience those effects[28]. These conditions erode economic stability and undermine coping mechanisms, thereby increasing susceptibility to mental health problems. In addition, displacement is frequently accompanied by food insecurity, inadequate shelter, and limited access to healthcare services[38,39]. These stressors, combined with the previously observed associations between food insufficiency and depression, likely compound the mental health burden among IDPs.

Taken together, these findings highlight the cumulative effect of economic loss, food insecurity, and forced displacement in elevating the risk of antenatal depression.

Exposure to conflict-related traumatic events was associated with significantly higher odds of antenatal depression (AOR = 2.3) compared with those who had not experienced such events. Evidence from systematic reviews conducted by the World Health Organization Regional Office for the Eastern Mediterranean indicates that women living in fragile and conflict-affected settings experience disproportionately higher levels of depression[40]. Furthermore, evidence from post-conflict settings, such as a study conducted seven years after the Northern Uganda conflict, demonstrates that cumulative exposure to war-related traumatic events is strongly associated with increased odds of major depressive disorder[36]. A more recent study from Northeast Ethiopia has similarly reported elevated odds of depression among war-affected populations, particularly among individuals with coexisting post-traumatic stress disorder (Post-traumatic stress disorder) and those perceiving high levels of life threat during conflict[35]. In addition, research conducted in internally displaced persons (IDP) camps in Northwest Ethiopia has identified female sex, witnessing killings, and direct exposure to traumatic life events as factors associated with increased odds of depression[41]. Taken together, these findings suggest that both the intensity and cumulative burden of traumatic experiences during conflict contribute to the increased risk of antenatal depression.

Poor partner relationship quality was significantly associated with higher odds of depression (AOR = 2.12) compared with good relationship quality. This finding is consistent with evidence from southern Ethiopia, where good husband support was protective against antenatal depression (AOR = 0.40), and from Babile district in eastern Ethiopia, where marital dissatisfaction (AOR = 2.37) and partner conflict in Gida Ayana district (AOR = 3.49) were associated with increased odds of depression [17,18,34]. Taken together, these findings suggest that the quality of intimate partner relationships plays an important role in maternal mental health, with supportive partnerships potentially buffering psychological distress, while conflictual or unsatisfactory relationships may increase vulnerability to depression.

Having less perceived neighborhood safety is associated with increased odds of depression (AOR: 2.58) compared to having high perceived neighborhood safety, as measured using the perceived neighborhood safety scale as adapted from the multi-ethnic atherosclerosis study (MESA). This finding aligns with increased new evidence suggesting the association between individual perception of neighborhood safety and mental health outcomes, particularly depression. For instance, a large population-based study in the Netherlands found that perceived neighborhood safety is inversely associated with depression severity, as measured using PHQ-9, the same tool used in the present study, to measure depression, highlighting the importance of subjective environmental perceptions in shaping mental health outcomes[42]. Similarly, a study conducted in low-income, predominantly African American populations in Detroit, USA, found a statistically significant inverse relationship between perceived neighborhood safety and depression scores[43].

Experiencing intimate partner violence (IPV), assessed using the Abuse Assessment Screen, was significantly associated with higher odds of depression (AOR = 2.25). This finding is consistent with national and international evidence, including an Ethiopian systematic review and meta-analysis of 30 studies showing increased odds of depression among women experiencing IPV (AOR = 3.09) [1], and a global umbrella review of six reviews and 73 studies identifying current or previous abuse as a significant risk factor for antenatal depressive symptoms [4]. These findings reinforce IPV as a consistent determinant of maternal depression, potentially with greater implications in conflict-affected settings.

Collectively, these findings indicate that antenatal depression in post-conflict Shire is influenced by interconnected individual, socioeconomic, relational, environmental, and conflict-related factors. Accordingly, prevention strategies should extend beyond routine antenatal care to integrate psychosocial support, economic strengthening, violence prevention, and interventions addressing the consequences of conflict and displacement.

### Strengths and limitations

This study provides context-specific evidence on antenatal depression in post-war and displacement-affected Shire, where empirical evidence remains limited. Inclusion of both internally displaced and host-community women enhances understanding of mental health disparities in fragile settings. The use of the validated PHQ- 9 and standardized measures, including the Abuse Assessment Screen and adapted MESA measures, strengthened outcome and exposure assessment. Furthermore, assessing determinants across individual, interpersonal, institutional, and community levels provided a comprehensive perspective on factors associated with antenatal depression.

Several limitations should be considered. The facility-based design, restricted to women attending antenatal care, may limit generalizability, particularly to women with limited access to health services. Retrospective assessment of pre-war economic status and conflict-related experiences may have introduced recall or misclassification bias, while responses to sensitive topics such as intimate partner violence and relationship quality may have been affected by social desirability bias. Residual confounding from unmeasured factors cannot be excluded; notably, social support was omitted because of a coding error affecting the Oslo Social Support Scale (OSS-3) in KoboToolbox. The perceived neighbourhood safety scale also showed relatively low internal consistency (Cronbach’s α = 0.57; McDonald’s ω = 0.61), potentially reducing measurement precision. Finally, the cross-sectional design precludes causal inference and does not rule out reverse causation, whereby depressive symptoms may have influenced perceptions of relationship quality or neighbourhood safety.

## Conclusion

This study found a substantial prevalence of antenatal depression (34.3%) among pregnant women attending public health centers in Shire, with an even greater burden among internally displaced women. Multivariable analysis identified independent determinants across individual, interpersonal, and community levels, including low income, unintended pregnancy, multiparity, poor pre-war economic conditions, conflict-related experiences, intimate partner violence, poor partner relationship quality, and low perceived neighborhood safety, while higher income was protective. These findings highlight antenatal depression as a major public health concern in this post-war, displacement-affected setting and underscore the need for integrated interventions addressing economic vulnerability, war-related experiences, interpersonal relationships, and community safety.

## Data Availability

The corresponding author will provide any data upon request.

## Acknowledgements

We sincerely thank Aksum University’s College of Health Sciences and the Comprehensive Specialized Hospital for their academic mentorship and institutional backing throughout this research. We are also deeply grateful to the Shire Wereda Health Office for supplying valuable background information on the city, which helped ground the study in its local context. Finally, we thank the data collectors, supervisors, study participants, and participating health centers for their commitment and essential contributions to the data collection process.

## List of abbreviations and acronyms

ANC: Antenatal Care
AND: Antenatal Depression
AOR: Adjusted Odds Ratio
BDI: Beck Depression Inventory
CI: Confidence Interval
COR: Crude Odds Ratio
DM: Diabetes Mellitus
EDRMC: Ethiopian Disaster Risk Management Commission
EPDS: Edinburgh Postnatal Depression Scale
FAHC: Five Angels Health Center
GBV: Gender-Based Violence
HIV: Human Immunodeficiency Virus
IDPs: Internally Displaced Persons
IPV: Intimate Partner Violence
SDG: Sustainable Development Goals
UHC: Umer Health Centre
VIF: Variance Inflation Factor
WHO: World Health Organization

## Declarations

## Ethics approval and consent to participate

Ethical approval was granted by Aksum University College of Health Science (IRB No. 056/2026) with supporting letters from the Tigray Regional Health Bureau. The permission letter was submitted to the Shire Woreda Health Office and shared with the selected health centers.

All participants gave written informed consent after receiving a full explanation of the study’s aims, procedures, and risks/benefits, and were informed of their right to withdraw at any time without consequence. Confidentiality was maintained throughout: no personal identifiers were collected, and data were handled anonymously on encrypted, password-protected devices.

Participants disclosing recent IPV, GBV, or suicidal ideation were referred to local psychological first aid and support services (May Dimu IDP Center, Adi Kentibay, Adiwenfito, and Embadanso), with those needing clinical or psychiatric care referred to Five Angels Health Center and Suhul General Hospital.

## Consent for publication

Not applicable

## Availability of data and materials

This study is original, with full citation of sources. Data supporting the findings can be obtained from the corresponding author upon a reasonable request.

## Author’s contributions

DBG was the principal author, participating in the conceptualization, design, acquisition, analysis, and interpretation of the data; drafting the manuscript; and serving as the corresponding author. TB was the primary academic advisor, contributed for design, acquisition, analysis, and interpretation of the data, and critically revised the manuscript. TGH, WTG and YT had contributed for design, acquisition, analysis, and interpretation of the data and critically revised the manuscript for important intellectual content. All authors read and approved the final manuscript.

## References

1. Rtbey G, Andualem F, Nakie G, Takelle GM, Mihertabe M, Fentahun S, et al. Perinatal depression and associated factors in Ethiopia: a systematic review and meta-analysis. BMC Psychiatry. 2024;24: 822. doi:10.1186/s12888-024-06246-5

2. WHO. Mental Health, Brain Health and Substance Use. In: Perinatal mental health [Internet]. 25 May 2026 [cited 25 Apr 2026]. Available: https://www.who.int/teams/mental-health-and-substance-use/promotion-prevention/maternal-mental-health

3. Al-Mutawtah M, Campbell E, Kubis H-P, Erjavec M. Women’s experiences of social support during pregnancy: a qualitative systematic review. BMC Pregnancy Childbirth. 2023;23: 782. doi:10.1186/s12884-023-06089-0

4. Dadi AF, Miller ER, Bisetegn TA, Mwanri L. Global burden of antenatal depression and its association with adverse birth outcomes: an umbrella review. BMC Public Health. 2020;20: 173. doi:10.1186/s12889-020-8293-9

5. Abebe M, Asgedom YS, Gebrekidan AY, Tebeje TM. Antenatal depression among pregnant women in Ethiopia: An umbrella review. PLOS ONE. 2025;20: e0315994. doi:10.1371/journal.pone.0315994

6. Gelaye B, Rondon MB, Araya R, Williams MA. Epidemiology of maternal depression, risk factors, and child outcomes in low-income and middle-income countries. Lancet Psychiatry. 2016;3: 973–982. doi:10.1016/S2215-0366(16)30284-X

7. Dadi AF, Wolde HF, Baraki AG, Akalu TY. Epidemiology of antenatal depression in Africa: a systematic review and meta-analysis. BMC Pregnancy Childbirth. 2020;20: 251. doi:10.1186/s12884-020-02929-5

8. American psychatric Association. Diagnostic and Statistical Manual of Mental Disorders. 5th ed. 2022.

9. Bauer A, Knapp M, Alvi M, Chaudhry N, Gregoire A, Malik A, et al. Economic costs of perinatal depression and anxiety in a lower middle income country: Pakistan. J Affect Disord. 2024;357: 60–67. doi:10.1016/j.jad.2024.04.061

10. Cleary S, Orangi S, Garman E, Tabani H, Schneider M, Lund C. Economic burden of maternal depression among women with a low income in Cape Town, South Africa. BJPsych Open. 2020/04/03 ed. 2020;6: e36. doi:10.1192/bjo.2020.15

11. Jahan N, Went TR, Sultan W, Sapkota A, Khurshid H, Qureshi IA, et al. Untreated Depression During Pregnancy and Its Effect on Pregnancy Outcomes: A Systematic Review. Cureus. 2021;13: e17251. doi:10.7759/cureus.17251

12. Roddy Mitchell A, Gordon H, Lindquist A, Walker SP, Homer CSE, Middleton A, et al. Prevalence of Perinatal Depression in Low- and Middle-Income Countries: A Systematic Review and Meta-analysis. JAMA Psychiatry. 2023;80: 425–431. doi:10.1001/jamapsychiatry.2023.0069

13. Hanif S, Momo J-E-T, Jahan F, Goldberg L, Herbert N, Yeamin A, et al. Flooding and elevated prenatal depression in rural Bangladesh: A mixed methods study. PLOS Glob Public Health. 2025;5: e0004792. doi:10.1371/journal.pgph.0004792

14. Dadi AF, Akalu TY, Wolde HF, Baraki AG. Effect of perinatal depression on birth and infant health outcomes: a systematic review and meta-analysis of observational studies from Africa. Arch Public Health. 2022;80: 34. doi:10.1186/s13690-022-00792-8

15. Zhang L, Li P, Ge Q, Sun Z, Cai J, Xiao C, et al. Maternal Prenatal Depressive Symptoms and Fetal Growth During the Critical Rapid Growth Stage. JAMA Netw Open. 2023;6: e2346018. doi:10.1001/jamanetworkopen.2023.46018

16. Fisher J, Cabral de Mello M, Patel V, Rahman A, Tran T, Holton S, et al. Prevalence and determinants of common perinatal mental disorders in women in low- and lower-middle-income countries: a systematic review. Bull World Health Organ. 2012;90: 139G–149G. doi:10.2471/BLT.11.091850

17. Oljira L, Abdissa E, Lema M, Merdassa E, Feyisa JW, Desalegn M. Antenatal depression and associated factors among pregnant women attending antenatal care at public health facilities in the Gida Ayana district, Oromia Region, West Ethiopia, in 2022. Front Public Health. 2023;11. doi:10.3389/fpubh.2023.1176703

18. Borie YA, Siyoum M, Tsega A, Anbese G. Maternal Depression and Associated Factors Among Pregnant Women Attending Ante Natal Care, Southern Ethiopia: Cross-Sectional Study. Front Public Health. 2022;10. doi:10.3389/fpubh.2022.848909

19. Takelle GM, Nakie G, Rtbey G, Melkam M. Depressive symptoms and associated factors among pregnant women attending antenatal care at Comprehensive Specialized Hospitals in Northwest Ethiopia, 2022: an institution-based cross-sectional study. Front Psychiatry. 2023;14. doi:10.3389/fpsyt.2023.1148638

20. Tesfaye Y, Agenagnew L. Antenatal Depression and Associated Factors among Pregnant Women Attending Antenatal Care Service in Kochi Health Center, Jimma Town, Ethiopia. J Pregnancy. 2021;2021: 5047432. doi:10.1155/2021/5047432

21. Beketie ED, Kahsay HB, Nigussie FG, Tafese WT. Magnitude and associated factors of antenatal depression among mothers attending antenatal care in Arba Minch town, Ethiopia, 2018. PLOS ONE. 2021;16: e0260691. doi:10.1371/journal.pone.0260691

22. Biratu A, Haile D. Prevalence of antenatal depression and associated factors among pregnant women in Addis Ababa, Ethiopia: a cross-sectional study. Reprod Health. 2015;12: 99. doi:10.1186/s12978-015-0092-x

23. Bitew T, Hanlon C, Medhin G, Fekadu A. Antenatal predictors of incident and persistent postnatal depressive symptoms in rural Ethiopia: a population-based prospective study. Reprod Health. 2019;16: 28. doi:10.1186/s12978-019-0690-0

24. Ayele TA, Azale T, Alemu K, Abdissa Z, Mulat H, Fekadu A. Prevalence and Associated Factors of Antenatal Depression among Women Attending Antenatal Care Service at Gondar University Hospital, Northwest Ethiopia. PloS One. 2016;11: e0155125. doi:10.1371/journal.pone.0155125

25. Mossie TB, Sibhatu AK, Dargie A, Ayele AD. Prevalence of Antenatal Depressive Symptoms and Associated Factors among Pregnant Women in Maichew, North Ethiopia: An Institution Based Study. Ethiop J Health Sci. 2017;27: 59–66. doi:10.4314/ejhs.v27i1.8

26. Adhana M, Teka H, Gebremihael MW, Gebretnsae H, Legesse A, Abreha G, et al. Armed conflict and maternal health service utilization in Ethiopia’s Tigray Region: a community-based survey. BMC Public Health. 2024;24. doi:10.1186/s12889-024-20314-1

27. Krupelnytska L, Vavilova A, Yatsenko N, Chrzan-Dętkoś M, Morozova-Larina O, Uka A, et al. War in Ukraine vs. Motherhood: Mental health self-perceptions of relocated pregnant women and new mothers. BMC Pregnancy Childbirth. 2025;25: 253. doi:10.1186/s12884-025-07346-0

28. Gebreyesus A, Niguse AT, Shishay F, Mamo L, Gebremedhin T, Tsegay K, et al. Prevalence of depression and associated factors among community hosted internally displaced people of Tigray; during war and siege. BMC Psychiatry. 2024;24: 3. doi:10.1186/s12888-023-05333-3

29. Department of Economic and Social Affairs, Sustainable Development. Transforming our world: the 2030 Agenda for Sustainable Development. In: United Nations [Internet]. Available: https://sdgs.un.org/2030agenda

30. Union A. Press Release on the Cessation of Hostilities Agreement Between the Go vernment of the Federal Democratic Republic of Ethiopia (FDRE) and the Tigray Peoples’ Liberation Front (TPLF), 2 November 2022, Pretoria, south Africa. Afr Union Comm. 2022. Available: https://papsrepository.africanunion.org/handle/123456789/1751

31. Kasujja M, Omara S, Senkungu N, Ndibuuza S, Kirabira J, Ibe U, et al. Factors associated with antenatal depression among women attending antenatal care at Mubende Regional Referral Hospital: a cross-sectional study. BMC Womens Health. 2024;24: 195. doi:10.1186/s12905-024-03031-0

32. Heissler K, Porter C. Know Your Place: Ethiopian Children’s Contributions to the Household Economy. Eur J Dev Res. 2013;25. doi:10.1057/ejdr.2013.22

33. Abebe T. Changing Livelihoods, Changing Childhoods: Patterns of Children’s Work in Rural Southern Ethiopia. Child Geogr. 2007;5: 77–93. doi:10.1080/14733280601108205

34. Ahmed SJ, Merid M, Edessa D, Usso AA, Adem HA, Tariku M, et al. Prenatal depression among pregnant women attending public health facilities in Babile district, Eastern Ethiopia: a cross-sectional study. BMC Psychiatry. 2024;24: 339. doi:10.1186/s12888-024-05732-0

35. Anbesaw T, Kassa MA, Yimam W, Kassaw AB, Belete M, Abera A, et al. Factors associated with depression among war-affected population in Northeast, Ethiopia. BMC Psychiatry. 2024;24: 376. doi:10.1186/s12888-024-05812-1

36. Mugisha J, Muyinda H, Malamba S, Kinyanda E. Major depressive disorder seven years after the conflict in northern Uganda: burden, risk factors and impact on outcomes (The Wayo-Nero Study). BMC Psychiatry. 2015;15: 48. doi:10.1186/s12888-015-0423-z

37. Stover AD, Shulkin J, Lac A, Rapp T. A meta-analysis of cognitive reappraisal and personal resilience. Clin Psychol Rev. 2024;110: 102428. doi:10.1016/j.cpr.2024.102428

38. WHO. Mental health in emergencies. 5 June 2025 [cited 5 Mar 2026]. Available: https://www.who.int/news-room/fact-sheets/detail/mental-health-in-emergencies

39. Almeida LM, Moutinho AR, Siciliano F, Leite J, Caldas JP. Maternal Mental Health in Refugees and Migrants: a Comprehensive Systematic Review. J Int Migr Integr. 2024;25: 209–222. doi:10.1007/s12134-023-01071-3

40. Rabbani F, Zahidie A, Siddiqui A, Shah S, Merali Z, Saeed K, et al. A systematic review of mental health of women in fragile and humanitarian settings of the Eastern Mediterranean Region. East Mediterr Health J. 2024;30: 369–379. doi:10.26719/2024.30.5.369

41. Tadesse G, Nakie G, Fentahun S, Andualem F, Tinsae T, Kibralew G, et al. Depressive symptoms and correlations among war-survivor internally displaced persons in two IDP camps in northwest Ethiopia: a cross-sectional survey. BMC Public Health. 2025;25: 1007. doi:10.1186/s12889-025-22232-2

42. Helbich M, Hagenauer J, Roberts H. Relative importance of perceived physical and social neighborhood characteristics for depression: a machine learning approach. Soc Psychiatry Psychiatr Epidemiol. 2020;55: 599–610. doi:10.1007/s00127-019-01808-5

43. Pearson AL, Clevenger KA, Horton TH, Gardiner JC, Asana V, Dougherty BV, et al. Feelings of safety during daytime walking: associations with mental health, physical activity and cardiometabolic health in high vacancy, low-income neighborhoods in Detroit, Michigan. Int J Health Geogr. 2021;20: 19. doi:10.1186/s12942-021-00271-3

